# The symptomatology of fever: a step towards qualitative definition of fever

**DOI:** 10.1101/2021.08.03.21261558

**Authors:** Mayank Kapoor, Nitin Kumar, Prasan Kumar Panda

## Abstract

**Background:** The old definitions of fever are based on cross-sectional surveys of the population without analyzing the associated symptomatology.

**Objectives:** To analyze associated symptoms with fever in a longitudinal follow-up study.

**Methods:** In a longitudinal study over one year, 196 participants recorded three temperature readings daily, one after waking up, one between 12-3 PM, one before sleeping, and filled the symptomatology questionnaire in a thermometry diary.

**Results:** Per protocol analysis was done for febrile participants (n=144). Fatigue (50.3%), warmth (47.3%), headache/head heaviness (47.0%), feeling malaise/general weakness (46.7%), loss of appetite (46.5%), muscle cramps/muscle aches (45.6%), chills/shivering (44.6%), increased sweating (43.0%), nausea (42.5%), irritability (38.9%), increased breathing rate (37.1%), and restlessness/anxiety/palpitations (36.5%) were the symptoms maximally seen during the febrile phase. A higher number of associated symptoms are associated with higher temperature readings. Dehydration suggested the numerically highest temperature values (100.86 ± 1.70°F) but seen in few febrile patients.

**Conclusions:** Incorporation of symptom analysis in febrile patients is the need of the hour. Fatigue and warmth are found to be most prevelent symptoms during febrile phase. Associated symptoms can help in predicting the intensity of fever also.

## Introduction

Fever has long been regarded as an important vital sign. Humans maintain their standard body temperature in a narrow range for the smooth functioning of the body.^1^ According to Wunderlich, the normal body temperature was defined as 37°C (98.6°F).^2^ His definitions were based on a cross-sectional study design, and there was no record of the associated symptoms of fever or complications. This is important, as the rise of temperature may be asymptomatic, as seen in the females in their menstrual cycle.^3^ Moreover, the definition of fever should include both quantitative and qualitative aspects as fever is a measurable sign, not an isolated symptom. However, no prior study has identified associated symptoms of fever for qualitative definition purpose.

The ideal way to define fever is to follow up healthy subjects prospectively. And when such individuals develop rise in baseline temperature with associated symptoms, fever is set to begin. Thereby, in the study, we recruited healthy participants and longitudinally followed them up and analyzed their temperature in association with their symptoms that were present at that time. We analyzed which symptomatology suggested fever and whether there was any relationship between no of symptoms and the absolute temperature values.

## Methods

A longitudinal study was conducted in a tertiary care hospital from July 2019 to September 2020 after approval from Institute Ethical Committee, All India Institute of Medical Sciences (AIIMS), Rishikesh (No. 235/IEC/PGM/2019). A sample size of 192 was calculated based on a prior study done by Mackowiak et al, and the participants were included in the study.^2^ The participants were selected based on simple random sampling. Healthy health care providers, students, and their accompanying family members aged 4-100 were included. Participants with a history of any diseases such as infections or trauma within the last month, chronic illness, and recent vaccination history in the last six months were excluded, including post-partum females (up to 8 weeks).

The study was conducted in three phases: non-febrile, febrile, and post-febrile phase. The participants recorded their temperature readings in all three phases. Minimum three oral temperature readings were recorded daily, one after waking up (AM), one between 12-3 PM (AN), and one before sleeping (PM). Participants filled a questionnaire related to the symptomatology of fever along with every temperature recording in provided clinical thermometry diary (supplemental material 1). The subjective sensation of feeling feverish was defined as the febrile phase’s onset and confirmed with a change in baseline temperature value for a participant.

The febrile phase data were analyzed as per-protocol analysis (for those participants only who developed the febrile phase during the study) using Statistical Package for Social Sciences (SPSS) Version 23. The frequency distribution of the various symptoms associated with fever was analyzed. The association of absolute values of temperature with the various symptoms was also done. Taking confidence interval as 95%, a P-value <0.05 was taken as statistically significant.

## Results

### Basic characteristics

The study screened 350 participants, and 144 were included in the analysis. The mean age of the participants was 24.24 ± 5.92 years (8-58 years). 26% (56) were < 20 years, 72.1% (155) were in the age group 20-40 years, and 1.9%(4) were >40 years of age. 52.1 % (112) were males. Various symptoms were present during the febrile phase, arranged in descending order (Table 1).

**Table 1:**
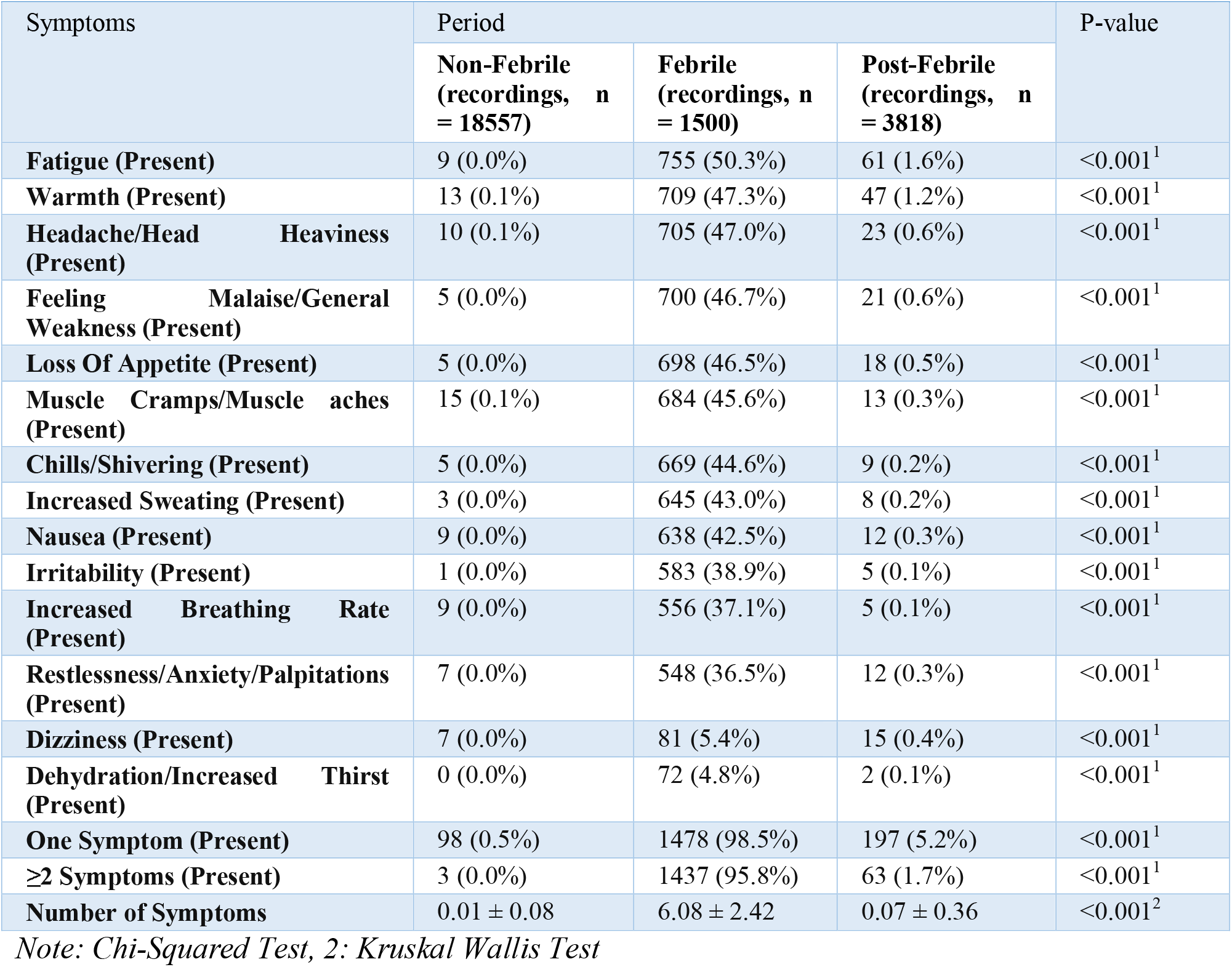
Symptomatology of three temperature phases among participants (n=144)

|  |  |  |  |  |
| --- | --- | --- | --- | --- |
| Symptoms | Period |  |  | P-value |
|  | Non-Febrile (recordings, n = 18557) | Febrile (recordings, n = 1500) | Post-Febrile (recordings, n = 3818) |  |
| **Fatigue (Present)** | 9 (0.0%) | 755 (50.3%) | 61 (1.6%) | <0.001<sup>1</sup> |
| **Warmth (Present)** | 13 (0.1%) | 709 (47.3%) | 47 (1.2%) | <0.001<sup>1</sup> |
| **Headache/Head Heaviness (Present)** | 10 (0.1%) | 705 (47.0%) | 23 (0.6%) | <0.001<sup>1</sup> |
| **Feeling Malaise/General Weakness (Present)** | 5 (0.0%) | 700 (46.7%) | 21 (0.6%) | <0.001<sup>1</sup> |
| **Loss Of Appetite (Present)** | 5 (0.0%) | 698 (46.5%) | 18 (0.5%) | <0.001<sup>1</sup> |
| **Muscle Cramps/Muscle aches (Present)** | 15 (0.1%) | 684 (45.6%) | 13 (0.3%) | <0.001<sup>1</sup> |
| **Chills/Shivering (Present)** | 5 (0.0%) | 669 (44.6%) | 9 (0.2%) | <0.001<sup>1</sup> |
| **Increased Sweating (Present)** | 3 (0.0%) | 645 (43.0%) | 8 (0.2%) | <0.001<sup>1</sup> |
| **Nausea (Present)** | 9 (0.0%) | 638 (42.5%) | 12 (0.3%) | <0.001<sup>1</sup> |
| **Irritability (Present)** | 1 (0.0%) | 583 (38.9%) | 5 (0.1%) | <0.001<sup>1</sup> |
| **Increased Breathing Rate (Present)** | 9 (0.0%) | 556 (37.1%) | 5 (0.1%) | <0.001<sup>1</sup> |
| **Restlessness/Anxiety/Palpitations (Present)** | 7 (0.0%) | 548 (36.5%) | 12 (0.3%) | <0.001<sup>1</sup> |
| **Dizziness (Present)** | 7 (0.0%) | 81 (5.4%) | 15 (0.4%) | <0.001<sup>1</sup> |
| **Dehydration/Increased Thirst (Present)** | 0 (0.0%) | 72 (4.8%) | 2 (0.1%) | <0.001<sup>1</sup> |
| **One Symptom (Present)** | 98 (0.5%) | 1478 (98.5%) | 197 (5.2%) | <0.001<sup>1</sup> |
| **≥2 Symptoms (Present)** | 3 (0.0%) | 1437 (95.8%) | 63 (1.7%) | <0.001<sup>1</sup> |
| **Number of Symptoms** | 0.01 ± 0.08 | 6.08 ± 2.42 | 0.07 ± 0.36 | <0.001<sup>2</sup> |

Note: Chi-Squared Test, 2: Kruskal Wallis Test

All symptoms were significantly present in the febrile phase. The mean temperature in the febrile phase was found to be 100.25 ± 1.44 ^0^F and various symptoms were associated with mean temperature (Table 2).

**Table 2:** Association between symptoms and temperature readings during febrile phase.

|  |  |  |
| --- | --- | --- |
| Symptoms | Temperature (°F) | P-value |
| **Chills/Shivering** |  | <0.001<sup>1</sup> |
| Present | 100.45 ± 1.43 |  |
| Absent | 98.07 ± 0.76 |  |
| **Dehydration/Increased Thirst** |  | <0.001<sup>1</sup> |
| Present | 100.86 ± 1.70 |  |
| Absent | 98.13 ± 0.86 |  |
| **Dizziness** |  | <0.001<sup>1</sup> |
| Present | 100.18 ± 1.87 |  |
| Absent | 98.13 ± 0.86 |  |
| **Fatigue** |  |  |
| Present | 100.25 ± 1.51 | <0.001<sup>1</sup> |
| Absent | 98.07 ± 0.74 |  |
| **Feeling Malaise/Generalized Weakness** |  |  |
| Present | 100.35 ± 1.44 | <0.001<sup>1</sup> |
| Absent | 98.07 ± 0.76 |  |
| **Headache/Head Heaviness** |  |  |
| Present | 100.26 ± 1.46 | <0.001<sup>1</sup> |
| Absent | 98.07 ± 0.76 |  |
| **Increased Breathing Rate** |  |  |
| Present | 100.61 ± 1.38 | <0.001<sup>1</sup> |
| Absent | 98.08 ± 0.77 |  |
| **Increased Sweating** |  |  |
| Present | 100.41 ± 1.41 | <0.001<sup>1</sup> |
| Absent | 98.08 ± 0.77 |  |
| **Irritability** |  |  |
| Present | 100.44 ± 1.46 | <0.001<sup>1</sup> |
| Absent | 98.08 ± 0.78 |  |
| **Loss Of Appetite** |  |  |
| Present | 100.26 ± 1.46 | <0.001<sup>1</sup> |
| Absent | 98.08 ± 0.77 |  |
| **Muscle Cramps/Muscle aches** |  |  |
| Present | 100.26 ± 1.48 | <0.001<sup>1</sup> |
| Absent | 98.08 ± 0.77 |  |
| **Nausea** |  |  |
| Present | 100.38 ± 1.46 | <0.001<sup>1</sup> |
| Absent | 98.08 ± 0.77 |  |
| **Restlessness/Anxiety/Palpitations** |  |  |
| Present | 100.50 ± 1.41 | <0.001<sup>1</sup> |
| Absent | 98.08 ± 0.78 |  |
| **Warmth** |  |  |
| Present | 100.25 ± 1.47 | <0.001<sup>1</sup> |
| Absent | 98.07 ± 0.76 |  |
| **One Symptom** |  |  |
| Present | 99.91 ± 1.55 | <0.001<sup>1</sup> |
| Absent | 98.00 ± 0.61 |  |
| **≥2 Symptoms** |  |  |
| Present | 100.22 ± 1.46 | <0.001<sup>1</sup> |
| Absent | 98.00 ± 0.61 |  |
| **Number of Symptoms** | Correlation Coefficient (rho) = 0.34 | <0.001<sup>2</sup> |

*Note: Wilcoxon-Mann-Whitney U Test, 2: Spearman Correlation*

## Discussion

We found that fatigue is the most common symptom associated with the febrile phase (50.3%), followed by warmth (47.3%). The febrile phase was associated with at least one symptom in 98.5% of the readings, whereas ≥ two were seen in 95.8%.The number of symptoms seen in various phases was: 0.01 ± 0.08 (non-febrile), 6.08 ± 2.42 (febrile), and 0.07 ± 0.36 (post-febrile) (P<0.001). One symptom was confirmly present when mean temperature was 99.91 ± 1.55°F, while mean temperature for two associated symptoms was 100.22 ± 1.46°F. As per our knowledge, no other study has analyzed the symptoms associated with fever.

As per classical teaching, fever is defined as an early morning temperature of >37.2°C (>98.9°F) or an evening temperature of >37.7°C (>99.9°F).^4^ The American College of Critical Care Medicine, the International Statistical Classification of Diseases, and the Infectious Diseases Society of America have defined fever as a temperature of ≥ 38.3 °C (100.9°F).^5^ However, no where it mentions about qualitative definition or associated symptoms since fever is a measurable sign. This is important, as the mere numerical rise in temperature can be seen physiologically also, as in females during the menstrual cycle.^3^ A study from British India demonstrated higher oral temperatures than those usually accepted as normal for temperate climates. The oral and rectal temperatures of Indian and British troops were higher than the normal range during the summer months.^6^ This again demonstrates that temperature is a quantitative variable undergoing fluctuations with the outside seasonal variations too. Hence qualitative definition is a must before any inference regarding the higher temperature range.

It is essential to derive a symptom-based definition of fever, where patients can be taught about a specific set of symptoms to look out for, suspecting fever. In the present scenario of COVID, it gathers more importance as due to the shortage of amenities, even thermometers cannot be ensured in every nook and corner. Hence, the patients can lookout for a few symptoms which may most likely predict fever. Present study confirms fatigue is most common associated symptoms (50.3%) during febrile phase followed by warmth (47.3%), headache/head heaviness (47.0%), feeling malaise/general weakness (46.7%), loss of appetite (46.5%), muscle cramps/muscle aches (45.6%), chills/shivering (44.6%), increased sweating (43.0%), nausea (42.5%), irritability (38.9%), increased breathing rate (37.1%), and restlessness/anxiety/palpitations (36.5%).

The presence of ≥ two symptoms was associated with higher temperature readings (100.22^0^F) compared to just one symptom (99.91^0^F). Similarly, dehydration correlated with higher temperature (100.86^0^F) followed by increase in breathing rate (100.61 ± 1.38 ^0^F). These findings can help in the early prediction of patients having a higher temperature and more likely to get sick. This may help in triaging the patients in the presently overburdened scenario of the COVID-19 pandemic also.

Our study has limitations too. Subjects were allowed to take antipyretics; hence, the symptoms could have resolved post drug use. The categorization amongst phases (febrile and non-febrile) was based on subjective parameters, varying from person to person. However, this itself forms the basis of our study as we propose that fever must be defined based on an individual’s assessment of the symptoms. Future study may establish objective measurements of these associated symptoms.

## Conclusion

Fever must not be defined as a mere rise of temperature, as this may occur physiologically also. As fever is a sign, various symptomatology of fever should be defined. Fatigue is found to be most prevelent symptoms (50.3%) during febrile phase followed by warmth, headache, malaise, loss of appetite, muscle aches, chills, increased sweating, nausea, irritability, increased breathing rate, and palpitations. Simultaneously higher rise in temperature is associated with more number of symptoms, thus predicting the intensity of fever.

## Data Availability

It will be made available to others as required upon requesting the corresponding author.

## Contributors

MK contributed to the data analysis and was involved in manuscript writing. NK contributed to the data collection and reviewing the manuscript. PKP gave the concept, interpreted analysis, critically reviewed the draft, and approved it for publication along with all authors.

## Data sharing

It will be made available to others as required upon requesting the corresponding author.

## Acknowledgment

Thanks to Dr Ajeet, Dr Minakshi, and Dr Yogesh for helping in protocol developement.

## References

1. Ganong’s Review of Medical Physiology, 26e | AccessMedicine | McGraw-Hill Medical. https://accessmedicine.mhmedical.com/book.aspx?bookID=2525. Accessed March 15, 2021.

2. Mackowiak PA, Wasserman SS, Levine MM. A Critical Appraisal of 98.6°F, the Upper Limit of the Normal Body Temperature, and Other Legacies of Carl Reinhold August Wunderlich. JAMA J Am Med Assoc. 1992;268(12):1578–1580. doi:10.1001/jama.1992.03490120092034

3. Steward K, Raja A. Physiology, Ovulation, Basal Body Temperature. StatPearls Publishing; 2019. http://www.ncbi.nlm.nih.gov/pubmed/31536292. Accessed March 28, 2021.

4. Harrison’s Principles of Internal Medicine, 20e | AccessMedicine | McGraw-Hill Medical. https://accessmedicine.mhmedical.com/book.aspx?bookID=2129. Accessed March 15, 2021.

5. Walter EJ, Hanna-Jumma S, Carraretto M, Forni L. The pathophysiological basis and consequences of fever. Crit Care. 2016;20(1):200. doi:10.1186/s13054-016-1375-5

6. Renbourn ET, Bonsall FF. Normal Body Temperatures in N. India. Vol 1. BMJ Publishing Group; 1946. https://www.ncbi.nlm.nih.gov/pmc/articles/PMC2059146/. Accessed March 15, 2021.

